# Serum BMP8B levels in women with polycystic ovary syndrome: a case-control study

**DOI:** 10.1101/2024.06.06.24308575

**Authors:** Hui-Ling Xue, Wan-Jiao Hao, Jie Song, Ye Li, Man Liu, Jun-XIU Wei, Xiao-Pang Ren

## Abstract

**Objectives:** Bone morphogenetic protein-8B (BMP8B) is an adipokine that is synthesized in many tissues and has been shown to be associated with the development of obesity and metabolic disorders in animals and humans. The aim of this study is to investigate the relationship between serum BMP8B levels and various metabolic parameters in women with PCOS.

**Material and methods:** This cross-sectional study included 80 women with PCOS and 40 age- and BMI-matched controls without PCOS. BMP8B, total cholesterol, triglyceride,fasting blood glucose (FBG), insulin, total testosterone (T),follicle stimulating hormone (FSH), luteinizing hormone (LH),estradiol (E2) levels were measured in all the participants. HOMA-IR was used to calculate the insulin resistance.

**Results:** Serum BMP8B levels were lower in women with PCOS than in healthy women(43.11±13.09 [ng/mL]vs. 106.45±52.32 [ng/mL], p< 0.001). HOMA-IR, LH/FSH, total-testosterone, total cholesterol, and triglyceride levels were significantly higher in women with PCOS than controls. Circulating BMP8B levels were positively correlated with HOMA-IR,BMI in PCOS group; was positively correlated with triglyceride in Control group.

**Conclusion:** The low concentration of circulating BMP8B in PCOS may be associated with insulin resistance,Lipid metabolism and BMI,bat may be a new target for PCOS treatment.

## INTRODUCTION

Polycystic ovary syndrome (PCOS) is the most common endocrinopathy affecting reproductive-aged women, with impacts across the lifespan from adolescence to post menopause. with a worldwide prevalence ranging from 6 to 21%, depending on the diagnostic criteria [1–5]., is characterized by ovulatory dysfunction, polycystic ovaries and biochemical (elevated androgens) and/or clinical (hirsutism, acne) hyperandrogenism. It is associated with insulin resistance, glucose intolerance, metabolic syndrome, obesity, hyperlipidemia, hypertension, chronic low-grade inflammation and increased risk of developing type 2 diabetes mellitus (T2DM) [6–9]. but the etiology of PCOS is unknown,the pathogenesis is complex, the clinical heterogeneity is large, the diagnostic criteria are still controversial, and the diagnosis and treatment choice and management strategy are different.

Brown adipose tissue(BAT) is a specialized fat tissue that serves as the primary site for adaptive nonshivering thermogenesis to generate heat under cold stress in mammals. BAT participates in primary metabolism and energy expenditure (EE), and it can be quickly stimulated by thermal or dietary stimulation [10]. A recent study showed that an increase in BAT mass and/or function could be an effective therapeutic target for the treatment of obesity and other related metabolic diseases in patients[11]. Brown adipokines are regulatory factors secreted by brown adipocytes that posses autocrine, paracrine, and endocrine activities and regulate BAT differentiation [12]. Some adipokines display hormonal functions that increase BAT activity, improve the metabolic profile of glucose and lipid homeostasis, and mediate the browning of WAT [13–15]. Bone morphogenetic protein-8B (BMP8B) is an adipokine produced by brown adipose tissue (BAT) contributing to thermoregulation and metabolic homeostasis in rodent models. [16]. Recent studies have shown that BMP8B plays an important role in energy metabolism and regulation of obesity. BMP8B can promote the increase of heat production, the acceleration of metabolism, and the reduction of body mass. It can also regulate lipid metabolism and has the effect of prosteatosis[17]. Yet, the role of BMP8B has not been fully understood in human homeostasis. Despite some studies on T2DM, metabolic syndrome and BMP8B, the relationship between BMP8B and PCOS has not been researched yet. For this reason, the aim of this study is to investigate the relationship between serum BMP8B levels and various metabolic parameters in patients with PCOS.

## MATERIAL AND METHODS

This cross-sectional clinical trial study was conducted in Affiliated Hospital of Hebei University. The institutional ethical committee has approved the trial protocol. We enrolled in the study a total of 120 patients presenting to our reproductive medicine clinics. The study group composed of 80 women diagnosed with PCOS while the control group consisted of age and BMI matched 40 women that had regular menstrual cycles. Detailed history of all patients was taken. After a physical exam we recorded anthropometric data (age, weight, height). Height [cm] and weight [kg] were measured with the patient barefoot in light daily clothes. BMI was calculated using the formula: weight [kg]/square meter of height [m2]. Patients were subdivided into obese, overweight,and control based on BMI: a BMI ≥28kg/m2 was considered obese, a BMI <24kg/m2 was considered control,the BMI between 24 and 28kg/m2 was overweight[18].Blood pressure was measured in the sitting position ensuring a min.of 15 [min] rest prior to measurement. The average of three measurements taken was calculated. The PCOS diagnosis was confirmed with at least two criteria according to the 2003 Rotterdam consensus: oligo-ovulation or anovulation, presence of polycystic ovaries on transvaginal ultrasound and hyperandrogenism present as a laboratory finding and/or symptoms. The biochemical hyperandrogenism was defined as total testosterone levels above the range that is regarded as normal (normal range: 0.01–0.48 nmol/L) [19]. For the evaluation of the polycystic ovaries’ (PCO) morphology, all women underwent transvaginal ultrasonography in the early follicular phase. The presence of PCO was diagnosed with the presence of 12 or more follicles with a diameter of 2–9 mm and/or increased ovarian volume (> 10 cm3) of each ovary. Menstrual periods were characterized by oligomenorrhea (absence of menstruation for 45 days or more) or by amenorrhea (no menstrual period for 3 months or more) in the PCOS group. The control group menstrual periods were regular and lasted 25–32 days. Additionally,the control group had neither hirsutism nor hyperandrogenism. All participants were non-smokers and did not have any medical treatment in the past. Pregnant or lactating women,hyperprolactinemia, Cushing’s syndrome, congenital adrenal hyperplasia, other diseases of the adrenal gland, thyroid disorders, impaired glucose tolerance, type 1 or type 2 diabetes mellitus and the use of insulin sensitizers or oral anti-diabetics were all excluded from the study.

### Laboratory studies

Antecubital venous blood samples were taken from each female in the early follicular phase of menstruation (2nd to 4th day). The samples were centrifuged and separated from the serum and stored at –80° C for analysis.

Total cholesterol, triglycerides, fasting blood glucose (FBG),serum insulin, total testosterone (T),follicle stimulating hormone (FSH), luteinizing hormone (LH),estradiol (E2) and BMP8B levels were measured.

Serum total cholesterol, and triglyceride levels were determined using an automated analyzer (Abbott Architect C 16000, IL, USA) with its own kits (Abbott Diagnostics, Wiesbaden, Germany).. Serum insulin levels were measured by an automated analyzer (Abbott Architect I2000, IL, USA) using a chemiluminescent microparticle immunoassay (CMIA) with its own kit (Abbott Diagnostics, Wiesbaden, Germany). Serum FSH, LH, E2, and T levels were measured with CMIA (Beckman Coulter Inc., Brea, CA, USA).Serum BMP8B concentrations were assessed by enzyme immunoassay (HENGYUAN, Shanghai,China).The intra-assay CV was < 10% and the inter-assay CV was < 12%. Using the formula stated for the homoeostasis model assessment of insulin resistance (HOMA-IR) method,the insulin resistance was calculated according to the formula:HOMA-IR = fasting insulin [mU/mL] × fasting glucose [mg/dL]/22.5.Patients with PCOS were grouped according to HOMA-IR: patients in the PCOS-IR group had HOMA-IR of ≥2.69 and those in the PCOS-non-IR group had HOMA-IR of <2.69[20,21].

### Statistical analysis

All statistical analysis was performed using the Statistical Package for the Social Sciences Software, version 18.0. The normality of the data was tested using the Kolmogorov-Smirnov test and all continuous variables had normal distribution (p > 0.05). Continuous variables were presented as mean ± standard deviation (SD). The demographic and laboratory characteristics of women with and without PCOS were compared using the two-tailed independ ent sample t-test. The relationship between BMP8B and other parameters was assessed by Pearson correlation analysis. To investigate the relationship between independent BMP8B levels in PCOS development, multiple linear regression analyzes were performed to adjust for equivalence and to determine the independent relationships of BMP8B levels with age, BMI,HOMA-IR, T, LH/FSH,E2, and triglyceride. All independent variables in the multiple linear regressions were tested for multicollinearity.If the variance inflation factor (VIF) exceeded 2.5, the variable was considered to be collinear. All reported confidence interval (CI) values are calculated at the 95% level. A two-sided p value of < 0.05 was considered statistically significant.

## RESULTS

1. Clinical and laboratory characteristics of the study population.The comparative demographic and laboratory parameters of women with and without PCOS are presented in Table 1. The cases were matched in terms of age. The parameter was similar and no statically significant differences were recorded between the groups (p > 0.05).Serum BMP8B levels were lower in women with PCOS than in healthy women and the difference was statically significant 43.11±13.09 ng/mL vs. 106.45±52.32ng/mL,p =0.000). Serum Triglyceride, LH, HOMA-IR, and T levels were all significantly higher in women with PCOS than in healthy women.
2. We divided PCOS patients into normal body weight group, overweight group and obesity group according to different BMI ( normal weight group 17 <BMI ≦ 24, overweight group 24<BMI ≦28, obesity group BMI>28), conducted inter-group comparison of each index, and compared it with the normal control group at the same time, so as to clarify the relationship between BMP8B and each index. Our results showed that, Patients with normal weight PCOS had higher levels of HOMA-IR, LH/FSHT, T and triglyceride than the normal control group with statistical significance, while BMP8B was significantly lower than the normal control group with statistical significance. Compared with the normal control group, overweight patients with PCOS had higher BMI, HOMA-IR, LH/FSHT, T and triglyceride, while BMP8B was significantly lower than the normal control group, which was statistically significant. Compared with the normal control group, the BMI, Total cholesterol, HOMA-IR, LH/FSHT, T and triglyceride of the obese patients with PCOS were all higher than the normal control group, while the BMP8B was significantly lower than the normal control group, which was statistically significant. The BMI, HOMA-IR and triglyceride levels of normal weight patients with PCOS were lower than those of overweight and obese patients with PCOS, which was statistically significant, while BMP8B was significantly higher than that of overweight and obese patients with PCOS, which was statistically significant. In overweight and obese patients with PCOS, there was no difference in all data except BMI.
3. We divided PCOS patients into insulin resistance group and non-insulin resistance group according to differences in HOMA-IR, and conducted inter-group comparison of all indicators, and compared them with normal control group at the same time, so as to clarify the relationship between BMP8B and the indicators. Our results show that, BMI, LH/FSHT, T, total cholestero and triglyceride were significantly higher in the PCOS non-insulin resistant group than in the normal control group. BMP8B decreased significantly. Compared to PCOS insulin resistance group There was no significant difference in other indexes except BMI and HOMA-IR. BMI, LH/FSHT, T, total cholestero and triglyceride were significantly higher in the PCOS insulin resistance group than in the normal control group. BMP8B decreased significantly.

**Table 1.** Comparison of demographic and laboratory characteristics of patients.

| Variables | Controls<br>n = 40 | PCOS<br>n = 80 | t | p |
| --- | --- | --- | --- | --- |
| Age (years) | 27.02±4.09 | 25.46±5.00 | 1.708 | 0.090 |
| BMI [kg/m<sup>2</sup>] | 21.53±2.83 | 28.18±5.18 | 7.560 | 0.000 |
| HOMA-IR | 2.08±0.53 | 5.00±3.14 | 5.814 | 0.000 |
| LH / FSH | 1.00±0.43 | 2.11±1.06 | 6.352 | 0.021 |
| T | 0.22±0.10 | 0.56±0.66 | 3.199 | 0.002 |
| BMP8B | 106.45±52.32 | 43.11±13.09 | 10.242 | 0.000 |
| Estradiol [pg/mL] | 39.00±13.66 | 38.12±16.57 | 0.288 | 0.774 |
| total cholesterol [mmol/L] | 4.27±0.61 | 4.64±0.84 | 0.289 | 0.015 |
| triglyceride [mmol/L] | 0.82±0.29 | 1.89±1.39 | 4.794 | 0.000 |

### 4. Correlation analysis

Pearson’s correlation coefficients are presented in Table 4. Correlation was calculated between BMP8B and various other parameters in the two groups of women. The triglyceride levels l were positively correlated with HOMA-IR, BMI and total cholesterol in the PCOS groups. Circulating BMP8B levels were negative correlation with BMI and HOMA-IR in the PCOS group.In the normal group, circulating BMP8B levels were significantly negatively correlated with triglycerides.

**Table 2.** PCOS was grouped according to BMI and the data were compared with each other.

| Variables | Controls<br>n = 40 | PCOS-normal weight<br>n = 18 | PCOS-overweight<br>n = 20 | PCOS-obese<br>n = 42 | P1 | P2 | P3 | P4 | P5 | P6 |
| --- | --- | --- | --- | --- | --- | --- | --- | --- | --- | --- |
| Age (years) | 27.02<br>±4.09 | 25.38<br>±5.32 | 25.35<br>±4.88 | 25.54<br>±5.04 | 0.206 | 0.167 | 0.151 | 0.981 | 0.913 | 0.885 |
| BMI [kg/m<sup>2</sup>] | 21.53<br>±2.83 | 21.26<br>±2.62 | 26.10<br>±1.02 | 32.15<br>±3.02 | 0.727 | 0.000 | 0.000 | 0.000 | 0.000 | 0.000 |
| HOMA-IR | 2.08 ± 0.53 | 2.88 ± 1.25 | 5.69 ± 4.03 | 5.58 ± 2.87 | 0.001 | 0.000 | 0.000 | 0.008 | 0.000 | 0.903 |
| LH / FSH | 1.00 ± 0.43 | 2.09 ± 1.08 | 2.03 ± 1.23 | 2.16 ± 0.99 | 0.000 | 0.000 | 0.000 | 0.874 | 0.788 | 0.638 |
| T[ng/mL] | 0.22 ± 0.10 | 0.84 ± 1.35 | 0.49 ± 0.24 | 0.48 ± 0.17 | 0.006 | 0.000 | 0.000 | 0.270 | 0.980 | 0.863 |
| BMP8B | 106.45 ± 52.32 | 53.31 ± 19.00 | 36.99 ± 3.92 | 41.65 ± 10.37 | 0.000 | 0.000 | 0.000 | 0.001 | 0.003 | 0.057 |
| Estradiol [pg/mL] | 39.00 ± 13.66 | 38.11 ± 18.75 | 41.88 ± 19.02 | 36.34 ± 14.34 | 0.840 | 0.502 | 0.393 | 0.542 | 0.691 | 0.206 |
| Total cholesterol [mmol/L] | 4.27 ± 0.61 | 4.45 ± 0.79 | 4.56 ± 1.13 | 4.77 ± 0.61 | 0.358 | 0.211 | 0.001 | 0.742 | 0.124 | 0.368 |
| triglyceride [mmol/L] | 0.82 ± 0.29 | 1.16 ± 0.48 | 1.71 ± 0.78 | 2.30 ± 1.72 | 0.002 | 0.000 | 0.000 | 0.015 | 0.008 | 0.151 |

P1:Controls.vs.PCOS- normal weight ;P2:Controls.vs. PCOS- overweight;P3:Controls.vs. PCOS- obese; P4: PCOS-normal weightvs. PCOS- overweight;P5: PCOS-normal weight.vs. PCOS- obese; P6: PCOS- overweight.vs. PCOS- obese。

**Table 3..** PCOS was grouped according to HOMA-IR and the data were compared with each other.

| Variables | Controls<br>n = 40 | PCOS-non-IR<br>n =19 | PCOS-IR<br>n = 61 | P1 | P2 | P3 |
| --- | --- | --- | --- | --- | --- | --- |
| Age (years) | 27.02±4.09 | 25.31±5.72 | 25.50±4.81 | 0.195 | 0.104 | 0.885 |
| BMI [kg/m2] | 21.53±2.83 | 24.48±5.6 | 29.34±4.48 | 0.010 | 0.000 | 0.000 |
| HOMA-IR | 2.08±0.53 | 2.21±0.48 | 5.87±3.11 | 0.396 | 0.000 | 0.000 |
| LH / FSH | 1.00±0.43 | 1.97±1.06 | 2.16±1.07 | 0.000 | 0.000 | 0.516 |
| T | 0.22±0.10 | 0.59±0.23 | 0.56±0.75 | 0.000 | 0.007 | 0.859 |
| BMP8B | 106.45±52.32 | 45.92±18.84 | 42.23±10.77 | 0.000 | 0.000 | 0.286 |
| Estradiol [pg/mL] | 39.00±13.66 | 39.27±21.58 | 37.76±14.85 | 0.953 | 0.675 | 0.732 |
| total cholesterol [mmol/L] | 4.27 ± 0.61 | 4.75 ± 0.82 | 4.61 ± 0.85 | 0.016 | 0.033 | 0.537 |
| triglyceride [mmol/L] | 0.82 ± 0.29 | 1.45 ± 0.67 | 2.03 ± 1.53 | 0.000 | 0.000 | 0.110 |

P1:control.vs.PCOS-NON-IR; P2:control.vs.PCOS-IR;P3 P3: PCOS-NON-IR. vs. PCOS-IR;

**Table 4.** Correlation coefficients between BMP8B levels and clinical parameters.

|  | BMP8B |  |  |  |
| --- | --- | --- | --- | --- |
|  | PCOS |  | Control |  |
|  | r | P | r | P |
| Age | -0.123 | 0.275 | -0.025 | 0.877 |
| BMI | -0.240 | 0.032 | -0.204 | 0.207 |
| HOMA-IR | -0.246 | 0.028 | 0.100 | 0.538 |
| LH / FSH | 0.061 | 0.592 | -0.111 | 0.931 |
| T | -0.138 | 0.224 | -0.047 | 0.772 |
| Estradiol | -0.024 | 0.832 | -0.157 | 0.335 |
| total cholesterol | 0.102 | 0.369 | 0.113 | 0.487 |
| triglyceride | -0.079 | 0.486 | -0.436 | 0.005 |

## DISCUSSION

The clinical features of PCOS, such as insulin resistance, obesity, dyslipidaemia and hyperandrogenism, can be classified as metabolic syndrome. Accordingly, 43% of adult women and nearly one-third of adolescent teenagers with PCOS have metabolic syndrome [22]. IR, obesity and hyperandrogenism are inseparable in the pathogenesis of PCOS. Insulin resistance (IR) is common in PCOS patients. IR has been reported in approximately 50–80% of women with different phenotypes of PCOS in different races [23,24,25]. Hyperinsulinaemia caused by IR exerts a cogonadotropin effect on the ovaries and decreases the expression of sex hormone-binding protein (SHBG), leading to the onset of hyperandrogenism [26,27]. Androgens can induce the accumulation of adipose tissue, especially abdominal fat tissue, and cause IR in subcutaneous adipose tissue [28,29]. Obesity, especially abdominal obesity,is associated with a variety of clinical features of PCOS. For example, due to adipose tissue dysfunction, adipocytes secrete non-physiological levels of adipokines, including IL6, IL8, TNF-α, leptin, adiponectin, resistin, lipocalin 2, monocyte chemoattractant protein-1 (MCP1), retinol binding protein-4 (RBP4), and CXC-chemokine ligand 5 (CXCL5), which may be involved in IR [30,31,32,33]. Obesity results from the proliferation of fat cells, three distinct stages can be divided for the process of adipogenesis: (1) commitment of MSCs to the lineage of adipocytes; (2) clonal expansion involving DNA and cell replication; (3) terminal differentiation, which involves the activation of specific transcription factors such as the peroxisome proliferator-activated receptor gamma (PPARγ) and the CCAAT/enhancer-binding protein (C/EBP) family. Changes in any of these three stages can affect fat formationIt has been reported that the prevalence of dyslipidaemia in PCOS patients is 70%, and the levels of low-density lipoprotein cholesterol (LDL-c), very-low-density lipoprotein cholesterol (VLDL-c), triglycerides (Tgs), and free fatty acid are increased, while the levels of high-density lipoprotein cholesterol (HDL-c) are decreased [34,35]. Dyslipidaemia is regarded as an important metabolic phenotype, although it is not a diagnostic criterion.

BAT was first described in small mammals and infants as an adaptation to defend against the cold [36,37]. BAT is the major site of adaptive non-shivering thermogenesis, both during cold exposure and after meals in so called postprandial thermogenesis. The inverse relationship between the BAT activity and body fatness suggests that BAT, through increasing energy expenditure, is protective against body fat accumulation [38,39] This has led to BAT being viewed as a promising therapeutic target for combating human obesity and related metabolic disorders [40,41]. In addition, BAT also has a certain secretory effect, as an endocrine organ, it can secrete regulatory molecules that may affect metabolism, called batokines. It can communicate signals between multiple tissues and organs and play a role in systemic metabolic regulation. The identification of these secretory factors acting on the whole body is expected to provide a theoretical basis and scientific basis for the treatment of obesity and related chronic metabolic diseases. It was found that adipokine production in brown adipocytes was more responsive to androgen therapy than in white adipocytes[42].Bone morphogenetic protein 8B (BMP8B), a member of the transforming growth factor beta (TGFβ)-BMP superfamily, has recently emerged as a new batokine, secreted by brown/beige adipocytes [43].Current data point to BMP8B as a major molecule in whole body metabolism by modulating i) BAT thermogenesis and the browning of white adipose tissue (WAT), through its action on AMP-activated protein kinase (AMPK) in the ventromedial nucleus of the hypothalamus[43,44,45].

In our study, it was found that the serum BMP8b level of patients with PCOS was significantly lower than that of the normal control group, with statistical significance, which may be related to the high androgen in patients with PCOS. Studies have shown that testosterone can reduce UCP1 and mitochondrial biogenesis in BAT in rats, inhibit the transcription of PGC1a, a key regulator of UCP1 expression and mitochondrial biogenesis, significantly damage mitochondrial function, reduce mitochondrial respiratory function, and inhibit BAT function [42,46,47]. In our further research, we found that; The serum BMP8b level of PCOS patients with insulin resistance is also significantly lower than that of PCOS patients with normal insulin secretion, which may be because the decreased mitochondrial respiratory activity is also implicated in part, by insulin resistance [48,49,50]. YAN et al. [51] found that after YTHDF1 gene was knocked out in male mice fed high fat, body weight increase, glucose intolerance and insulin resistance unrelated to feeding occurred, while mice with YTHDF1 overexpression showed significant remission of glucose intolerance and insulin resistance. The expression of BMP8B protein in YTHDF1 knockout mice was significantly reduced. The mechanism may be that YTHDF1 gene knockout reduces the 3’-terminal non-coding sequence1 luciferase activity of BMP8B and regulates the translation of BMP8B, suggesting that the decrease of BMP8B can lead to decreased glucose tolerance and insulin resistance in the body. Robinson and colleagues showed that while resting energy expenditure was similar in women with and without PCOS, postprandial thermogenesis was reduced in both obese and lean women with PCOS [52]. This decreased postprandial thermogenesis may predispose women with PCOS to weight gain and help to explain the increased prevalence of obesity in women with PCOS. In our study, the serum BMP8b level of overweight or obese PCOS patients was significantly lower than that of normal weight patients, which may be related to the effect of BMP8b on adipogenesis. we showed that BMP8B down-regulates transcriptional regulators PPARγ and C/EBPα, thereby impeding the differentiation of 3T3-L1 preadipocytes into fully mature adipocytes. BMP8B increased the phosphorylation levels of SMAD2/3, and TP0427736 HCl (SMAD2/3 inhibitor) significantly reduced the ability of BMP8B to inhibit adipocyte differentiation, suggesting that BMP8B repressed adipocyte differentiation through the SMAD2/3 pathway. Moreover, the knockdown of BMP I receptor ALK4 significantly reduced the inhibitory effect of BMP8B on adipogenesis, indicating that BMP8B triggers SMAD2/3 signaling to suppress adipogenesis via ALK4. In addition, BMP8B activated the NF-κB signal, which has been demonstrated to impede PPARγ expression. Collectively, our data demonstrated that BMP8B activates both SMAD2/3 and NF-κB signals to inhibit adipocyte differentiation[53,54],leads to a buildup of fat cells that can lead to overweight or obesity. In addition, we found that triglyceride level in PCOS patients was significantly higher than that in normal patients, which was negatively correlated with serum BMP8B level and positively correlated with BMI and IR of patients. This suggests that BMP8B is involved in the regulation of lipid metabolism. When the lipid metabolism in the body is abnormal, the precursor of androgen synthesis, cholesterol, will produce a large amount of androgens under the action of 17α-hydroxylase and 1720-lyase, while obese PCOS women have more serious endocrine hormone disorders and lipid metabolism abnormalities[55,56].

In summary, by studying the expression of brown adipokine BMP8B in the serum of PCOS patients and its relationship with PCOS, obesity, insulin resistance and triglyceride, this paper suggests that BMP8B can promote thermogenesis and weight loss in obesity. At the same time, BMP8B is involved in the regulation of body fat metabolism,can improve glucose intolerance and insulin resistance. In addition, studies have shown that stimulating BAT activity [57] or transplanting BAT into rodent models of PCOS can improve reproductive and metabolic function [58].Therefore, we speculate that BAT may be a new target for PCOS treatment.

## Data Availability

no

